# Clinician Performance in Training Data Curation for an Arrhythmia Machine Learning Model: Is Anyone Qualified?

**DOI:** 10.64898/2026.01.07.26343629

**Authors:** Michael E Kim, Azadeh Assadi, Daniel Ehrmann, Will Dixon, Spencer Vecile, Robert Greer, Sebastian Goodfellow, Anica Bulic

## Abstract

**Background:** Early identification of arrhythmias in the intensive care unit (ICU) is important to prevent ICU morbidity and mortality. Timely arrhythmia detection relies on bedside providers’ telemetry interpretation. Machine learning (ML) models can function as clinical support tools to facilitate diagnoses. ML model development requires well-curated training data. The differential performance between labelers of different roles and experience is currently unknown.

**Methods:** This was a prospective observational study with frontline providers. 300 (200 original, 100 duplicate) 10-second telemetry tracings were labeled including sinus rhythm, 2^nd^/3^rd^ degree atrioventricular (AV) block, junctional ectopic tachycardia (JET), ectopic atrial tachycardia (EAT), and reentrant supraventricular tachycardia (SVT). Interrater reliability was calculated against the ground truth label as the primary performance measure (intrarater reliability for consistency utilizing duplicate labels).

**Results:** 11 participants completed the study: 1 Cardiology fellow, 4 pediatric ICU fellows, 2 pediatric cardiac ICU fellows, 3 pediatric cardiac ICU NPs, and 1 Pediatrics resident. Highest level of agreement was moderate (κ 0.68, p <0.001) with the majority poor to moderate. There was no association of clinical subspecialty with labeling performance. Performance varied by rhythm type (median κ): sinus (0.61), AV block (0.68) > Junctional (0.47), EAT (0.25), SVT (0.49). There was good intrarater reliability (κ 0.71 [median], p<0.001).

**Conclusions:** Overall, frontline provider performance was poor especially for complex arrhythmia classes. Cardiology training and experience was not associated with better performance. These findings highlight the need for thoughtful consideration in labeler training and validates the need for a clinical decision support tool in arrhythmia detection.

## Introduction

Early identification of arrhythmias in the cardiac intensive care unit (ICU) is imperative in the prevention of significant patient morbidity and mortality.^1^ Bedside telemetry, 12-lead electrocardiograms (ECG), and continuous physiological data monitoring systems are commonly utilized in the ICU to facilitate timely recognition. All these forms of monitoring require a healthcare provider, whether it is the bedside nurse, frontline provider, or staff physician, to actively analyze and interpret changes.

A machine learning (ML) model can serve as a clinical decision support tool (CDS) by signaling rhythm changes and providing a predicted diagnosis.^2–4^ Previous studies have examined its utility in recognition of atrial fibrillation^5^ and to facilitate interpretation of 12-lead ECGs.^2–6^ However, development and training a ML model for arrhythmia detection requires accurately labelling a significant amount of rhythm data.^2–4^ As labelling resources are limited, teams interested in developing rhythm identification CDS tools commonly seek to expand their pool of data labelers, but it is unknown who is qualified to function as a labeler in this capacity.

We conducted a study amongst healthcare providers who actively assess and interpret bedside telemetry to understand if clinical expertise and subspecialty training are modifiers of labeling performance. We hypothesized that individuals with formal cardiology training or 3+ years of experience working in a cardiac intensive care unit would perform better (Cohen’s κ >0.8) than those with minimal exposure to rhythm interpretation (Cohen’s κ >0.6).

## Methods

This is a prospective observational study conducted at a large quaternary children’s hospital. The Heart Center at SickKids performs approximately 500 cases requiring cardiopulmonary bypass annually and the cardiac intensive care unit admits 800 patients annually. The study was reviewed and approved by the SickKids Research Ethics Board (#1000081644).

Participants eligible for the study included frontline healthcare providers including physician trainees (residents and clinical fellows) and nurse practitioners across various subspecialties: general pediatrics, pediatric cardiology, pediatric critical care, pediatric anesthesia, and pediatric cardiac critical care. Individuals were recruited by email and tasked with labelling 300 10-second, single lead (limb lead II) telemetry strips to simulate the standard duration of a 12- or 15-lead ECG. The waveform labels were not cleaned or curated, maintaining realism with bedside telemetry interpretation. Label options included sinus rhythm, atrioventricular (AV) block (2^nd^ or 3^rd^ degree), junctional rhythm, ectopic atrial tachycardia (EAT), and reentrant supraventricular tachycardia (SVT). We defined sinus rhythm to include sinus bradycardia, sinus tachycardia, and normal sinus rhythm. Junctional rhythm was not constrained by heart rate thus included both junctional rhythm and junctional ectopic tachycardia. EAT was defined as a narrow QRS complex tachycardia with P-wave morphology that was not suspected to be sinus in origin. Reentrant SVT was defined as a narrow QRS complex tachycardia with no discernable P-wave morphology, variability, and significantly elevated rate (>180-200 beats per minute based on calculated normal ranges).

Ground truth labels for each strip were provided by a staff electrophysiologist (AB). There was an even distribution of rhythm classifications within the dataset which included 200 unique labels (40 each) and 100 (20 each) redundant labels. Redundancy was included to assess intrarater reliability with the waveform datasets. Participants provided informed consent and were given one month to complete the study. Participants were financially compensated for their participation.

### Label Studio

Label Studio is an open-source data labeling interface for various data types, including waveforms. Rhythms can be labeled as a whole or segmented into distinct regions. For this study, we utilized AtriumDB, a high-performance waveform data warehouse which has been established at SickKids since 2016, to extract our telemetry readings for this study.^7^ These readings were already labeled by a staff electrophysiologist (AB). Rhythms were de-identified before upload to label studio and linked to original patient information via a unique SHA-256 hash only accessible by the study team. The five rhythm waveforms for this study are seen in **Figure 1**. The Label Studio software displayed the timestamped ECG and heart rate signals collected by Philips^©^ monitors (which were stored and pulled from AtriumDB). A “normal heart rate range” based on the patient’s age was also included (the age itself was not entered into the software and thus not be accessed by the labelers, **Supplement 1**). Participants received a 1-on-1 tutorial session using Label Studio with example waveforms in addition to receiving an instructional document for independent review (**Supplement 2**).

**Figure 1.**
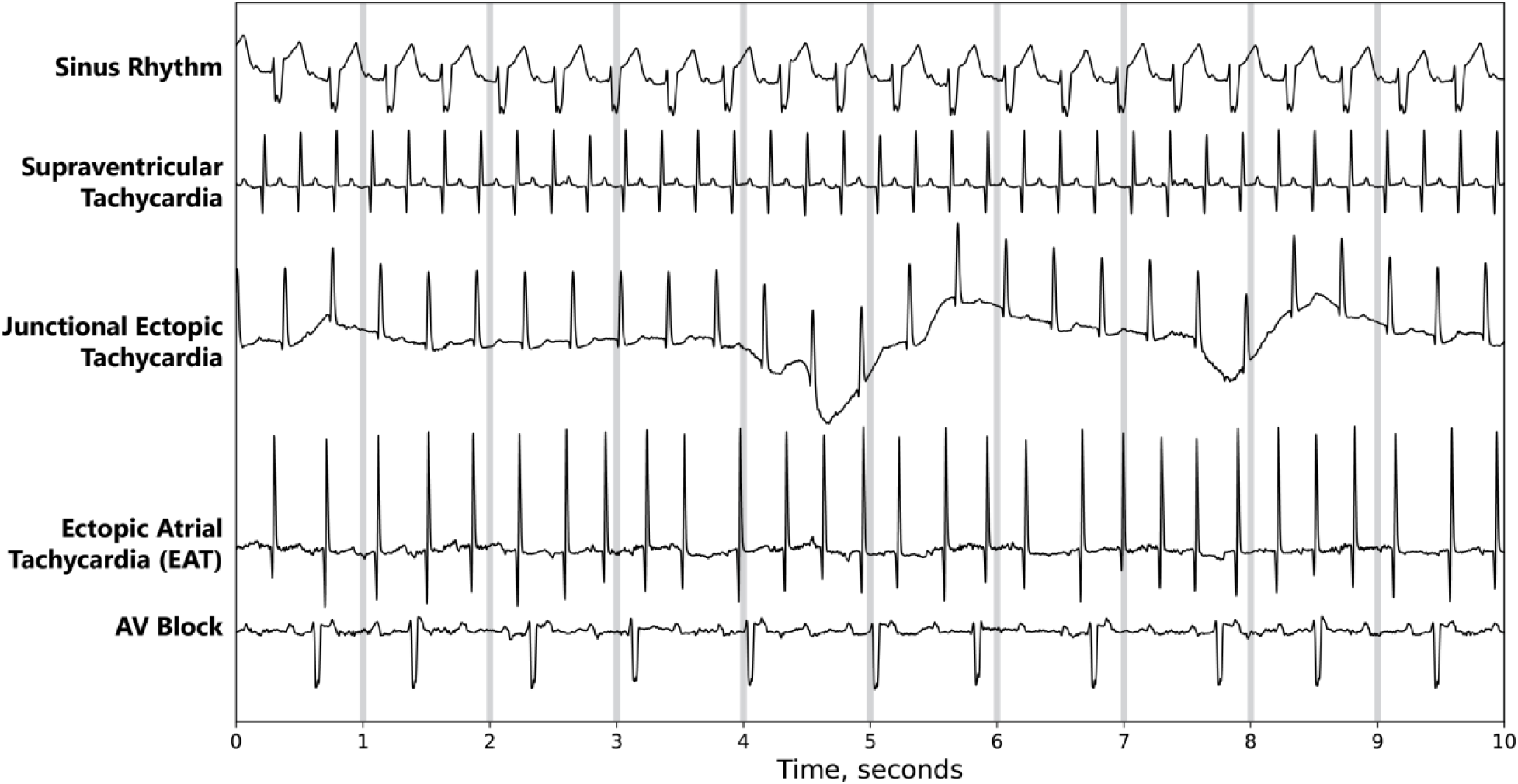
Rhythm waveform examples (five separate patients) that participants interpreted in Label Studio.

### Survey Design

Participants were asked to complete a survey at the end of the study (**Supplement 3**). The survey collected information regarding participant role, subspecialty, and years in practice. 4-point Likert-scale items addressed individual experience interpreting telemetry and comfort in doing so independently. Survey items were calculated by medians with interquartile ranges or frequencies with percentages, as appropriate.

### Statistical Methods

The sample size for the dataset was determined by the number of individual rhythm labels required for adequate power to calculate a Cohen’s kappa (κ) statistic in addition to considering study feasibility for participants to complete in a timely manner.^8^ We calculated a sample size (n=32) based on a two-rater system with an acceptable κ_0_ = 0.8, expected κ_1_ = 0.5, power = 0.8, and significance = 0.05. We increased this to 40 per rhythm class for a total of 200 and added 100 redundant readings to measure intrarater reliability (300 tasks combined). Individual performance with rhythm labeling was done utilizing Cohen’s κ computed against the ground truth label. The use of Cohen’s κ statistic with a “gold standard” comparator to participant labels has been accepted as a performance measure.^8,9^ Definitions for the level of interrater agreement in this study included: excellent (κ >0.9), very good (κ 0.8-0.89), good (κ 0.7-0.79), moderate (κ 0.6-0.69), and poor (κ <0.6). Intrarater reliability was calculated utilizing Cohen’s κ between the redundant labels within a given participant’s labelling session. Definitions for the level of intrarater agreement included: excellent (κ >0.8), good (κ 0.7-0.79), moderate (κ 0.6-0.69), and poor (κ <0.6).

## Results

### Demographics

There was a total of 11 participants in the study – 7 clinical fellows (1 pediatric cardiology, 2 pediatric critical care, 2 pediatric cardiac critical care, 2 pediatric anesthesiology), 3 cardiac intensive care nurse practitioners, and 1 general pediatrics resident.

### Labeling performance

Each participant labeled a set of 300 10-second single lead telemetry strips (**Table 1**). The level of agreement (Cohen’s κ) of labelers ranged from poor (0.37) to moderate (0.68) agreement with an average agreement of 0.5. When examining the interrater reliability further based on specific rhythm labels, there was high variability amongst participants with overall individuals having higher levels of agreement with sinus and AV block (**Table 2**). The remaining rhythm labels were poor overall with EAT having the lowest level of agreement. Intrarater reliability was overall good across all labelers (0.71) (**Table 1**).

**Table 1.** Inter- and intra-rater reliability amongst participants.

| **Labeler** | **Overall performance<sup>†</sup>** | **Intrarater reliability<sup>†</sup>** |
| --- | --- | --- |
| PEDS | 0.68 | 0.91 |
| CARD | 0.59 | 0.78 |
| CICU2 | 0.55 | 0.67 |
| CICU1 | 0.54 | 0.76 |
| NP1 | 0.53 | 0.71 |
| NP3 | 0.5 | 0.67 |
| PICU1 | 0.49 | 0.67 |
| ANE2 | 0.47 | 0.81 |
| PICU2 | 0.44 | 0.75 |
| ANE1 | 0.43 | 0.67 |
| NP2 | 0.37 | 0.46 |
| <sup>†</sup> expressed as Cohen's kappa ($\kappa$)<br>Abbreviations: PEDS – pediatrics resident; CARD – cardiology fellow; CICU – cardiac intensive care fellow; NP – nurse practitioner; PICU – pediatric critical care fellow; ANE – anesthesia staff | | |

**Table 2.** Interrater reliability separated by waveform label type.

| **Labeler** | **Sinus<sup>†</sup>** | **AV**<br>**block<sup>†</sup>** | **Junctional<sup>†</sup>** | **EAT<sup>†</sup>** | **SVT<sup>†</sup>** |
| --- | --- | --- | --- | --- | --- |
| CARD | 0.70 | 0.73 | 0.41 | 0.53 | 0.58 |
| ANE1 | 0.60 | 0.52 | 0.60 | 0.16 | 0.56 |
| ANE2 | 0.63 | 0.74 | 0.35 | 0.18 | 0.42 |
| PICU1 | 0.59 | 0.67 | 0.52 | 0.17 | 0.40 |
| PICU2 | 0.50 | 0.75 | 0.28 | 0.21 | 0.44 |
| NP1 | 0.62 | 0.68 | 0.51 | 0.33 | 0.49 |
| NP2 | 0.47 | 0.65 | 0.26 | 0.08 | 0.40 |
| NP3 | 0.59 | 0.64 | 0.47 | 0.25 | 0.58 |
| CICU1 | 0.66 | 0.68 | 0.47 | 0.50 | 0.38 |
| CICU2 | 0.61 | 0.65 | 0.58 | 0.25 | 0.59 |
| PEDS | 0.88 | 0.76 | 0.58 | 0.59 | 0.59 |
| <sup>†</sup>expressed as Cohen's kappa ($\kappa$)<br>Abbreviations: AV – atrioventricular; EAT – ectopic atrial tachycardia; SVT – supraventricular tachycardia; PEDS – pediatrics resident; CARD – cardiology fellow; CICU – cardiac intensive care fellow; NP – nurse practitioner; PICU – pediatric critical care fellow; ANE – anesthesia staff | | | | | |

We examined the performance of individual labelers based on sub-specialization, years of clinical experience, and self-described comfort with interpretation of bedside telemetry and 12-lead ECGs (**Figure 2**). There was no association with cardiology training or years of experience working in a cardiac unit with the level of agreement in the dataset. It is notable to mention that one individual who had <1 year of experience working with cardiac patients (a pediatric resident) had high scores for all rhythm classifications and overall performance. Participants with the lowest levels of agreement reported having lower confidence in independently classifying arrhythmias, clinical exposure to cardiac patients, or fundamental knowledge on electrophysiology (**Figure 2**).

**Figure 2.**
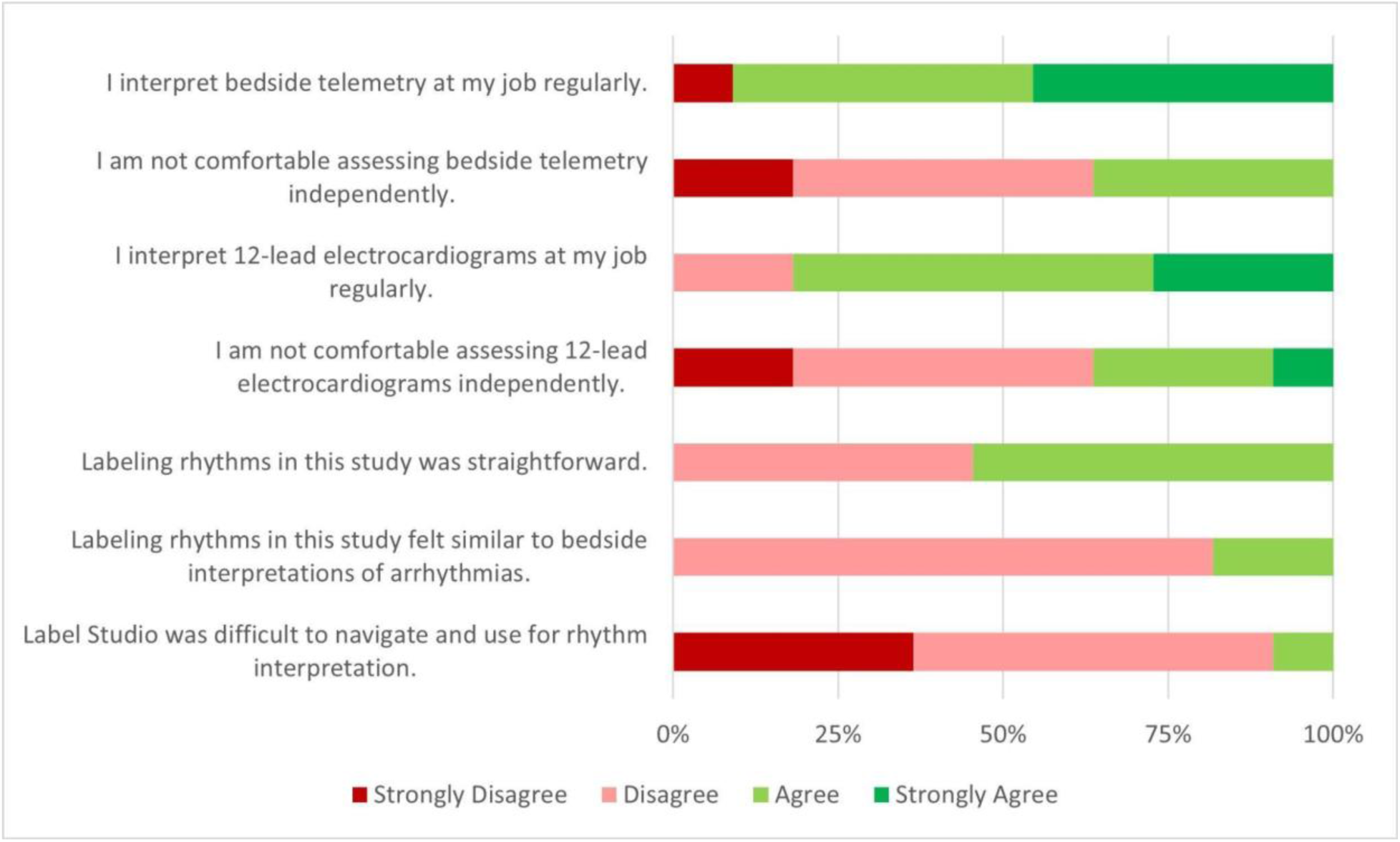
Likert-scale responses from participants regarding telemetry interpretation and Label Studio UI/UX.

### Survey Results

All participants completed the post-study survey (**Figure 1**). Overall, participants reported being responsible for interpreting bedside telemetry and 12-lead electrocardiograms, but at least half the group felt uncomfortable doing so independently. Individuals reported Label Studio was easy to utilize but found it to be significantly different from interpreting bedside telemetry. Most participants completed the labeling tasks within 4-8 hours on an average of 40 seconds – 1 minute per label (**Table 3**).

**Table 3.** Demographic information and labeling feasibility survey data.

| Item | n = 11 |
| --- | --- |
| **Clinical Role** |  |
| Clinical Fellow | 75 |
| Resident Physician | 1 |
| Nurse Practitioner | 3 |
| **Subspecialty** |  |
| Cardiology | 1 |
| Cardiac Intensive Care | 5 |
| Pediatric Critical Care | 2 |
| Pediatric Anesthesia | 2 |
| General Pediatrics | 1 |
| **Years in Clinical Practice** |  |
| <5 years | 2 |
| 5-10 years | 3 |
| >10 years | 6 |
| **Time to complete dataset** |  |
| <2 hours | 0 |
| 2-4 hours | 2 |
| 4-8 hours | 7 |
| >8 hours | 2 |
| **Time per label** |  |
| <1 minute | 6 |
| 1-5 minutes | 2 |
| >10 minutes | 0 |
| Variable (<1 minute to >10 minutes) | 3 |
| **Challenges with labeling** |  |
| User interface issues | 3 |
| Time (fatigue) | 5 |
| Waveform noise | 6 |
| Difficult to interpret | 6 |
| Other (specified) | 2 |
| No issues | 1 |
| **Participant reported rhythm labeling difficulty** |  |
| Sinus | 1 |
| Atrioventricular block | 1 |
| Junctional | 1 |
| Ectopic atrial tachycardia | 8 |
| Supraventricular tachycardia | 8 |
| None | 2 |

When asked about difficulties in labeling waveform data, individuals reported the main issues interpreting waveforms were the length of data provided (10 seconds was too short), significant noise, difficulty of rhythm diagnosis (without a 12-lead ECG), and labeling fatigue (**Table 3**). The most difficult rhythms to interpret and label included EAT and SVT as reported by participants.

## Discussion

Arrhythmia classification and labeling is a complex task where individual performance cannot rely solely on clinical experience. This was evident in our study sampling various frontline providers and determined overall performance of individuals ranged significantly from poor to moderate (depending on the rhythm class). There was no association between clinical subspecialty, role, or level of clinical experience with labeling performance. While participants were not informed of their performance, their responses in the survey reinforce the challenges of diagnosing arrhythmias based on a single waveform (lead II) with a fixed amount of data (10 seconds). While variance in knowledge, experience, and familiarity did not show dramatic increases in performance, another factor also includes the context in which these were completed (i.e. number of labels/session, attentiveness, diagnostic fatigue). These findings not only highlight the importance of training individuals who curate training data for a ML model but also validate the need for an arrhythmia CDS. In the ICU setting this becomes more impactful as several potentially life-threatening arrhythmias that were the hardest to diagnose in this study (junctional, EAT, SVT).

Arrhythmia detection and ML development have been studied since the late 1990s to early 2000s and are used routinely in 12-lead ECG analysis.^10,11^ While more sophisticated software architecture and coding now exist, an unavoidable roadblock to successful ML development includes the creation of a robust training dataset. Current research within the field has been mostly focused on adult studies which include recognition and prediction of atrial fibrillation, one of the most common arrhythmias within their population.^12^ While some studies have started to explore the potential of utilizing ECG waveform analysis for atrial fibrillation, there is a paucity of published models that explore more extensive ML arrhythmia detection models. This study highlights an important rate limiting step in the development process in that training data, especially for complex arrhythmias, requires trained labelers in which clinical experience alone may not be sufficient.

Curating a large training dataset of labeled heart rhythm data for CDS requires a corpus of labelers beyond pediatric electrophysiologists. There is uncertainty in determining what level of labeler performance is required to generate data sufficiently accurate to train a reliable ML model for arrythmia CDS. The first step in identifying adequate labeler performance is benchmarking relative to the gold standard, and this study sought to elucidate those benchmarks. We used realistic waveforms and evaluated frontline providers because they are exposed to arrythmias, have some training in rhythm diagnosis, and more numerous than subspecialty task physicians When comparing to the ground truth labels (those done by an electrophysiologist), we determined that it would be not feasible to have labeler qualifications to be restricted solely to those individuals. Since the problem of limited expert labelers persist, additional strategies should be considered. Additional study is needed to evaluate these strategies, which include training frontline providers (modules, workshops, etc.), hybrid labeling structures (e.g., first pass for frontline providers with verification of a random subset by experts), or other computational techniques (train on a subset of the data to build a model to help predict labels for more inexperienced labelers).

Consideration for future studies should include novel approaches in training data validation and creation. While clinical training alone is not indicative of the level of success in labeling, it would also be meaningful to determine if individuals without clinical or healthcare experience could be trained to recognize and label accurate data.

### Study Limitations

There are some limitations to acknowledge with this study. This was a small dataset including sample size of participants from a single center. Participants were from a mixed clinical background including those within the same specialty and/or clinical role which limits generalizability of performance based on those qualities alone. We believe this was an important first step in addressing this topic and would think there is benefit for future studies to expand both the size of the study group as well as the number of participating institutions. The study was not controlled in terms of how participants completed the study – some participants completed the dataset in one session versus others who did frequent daily sessions which can impact consistency and performance.

Another important consideration of this study is that all telemetry data was gathered from a single waveform (lead II) which can limit diagnostic utility for certain arrhythmias (i.e. EAT, SVT). The waveform data also were not filtered or curated for this study thus they did contain noise or variance – however these were all reviewed by our team before approving thus they were still diagnostically useful. It is also important to acknowledge labelers were given 10-second tasks which in comparison to the ground truth labels which were diagnosed based on a 2-minute task which could reveal more subtle findings (i.e. V-A dissociation in JET, rate variability for EAT). Finally, because we created an even number of rhythm types within the dataset, some labels were obtained from the same patient encounter and thus created additional redundant labeling tasks. Participants were not notified of this information, nor did their performance indicate as such.

Label Studio overall had favorable UI/UX feedback from a group that was taught to use the platform in a relatively short period of time. One important takeaway was that participants felt labeling rhythms were dissimilar to bedside telemetry interpretation which may also have impacted performance.

## Conclusion

This study sought to benchmark the labeling performance of individuals with varying clinical backgrounds. The overall level of agreement between labelers was lower than hypothesized. Rhythm difficulty, inexperience, lack of formal cardiology training, and perhaps most importantly lack of labeling experience may have contributed to our findings. At current state, frontline providers are not acceptable as the sole source for labeled pediatric heart rhythm data for CDS. Further study is warranted to understand the implications of these results on the ML-based arrhythmia detection CDS pipeline.

## Data Availability

All data produced in the present work are contained in the manuscript

## Abbreviations

ECG: electrocardiogram
ML: machine learning
AV: atrioventricular
EAT: ectopic atrial tachycardia
SVT: supraventricular tachycardia

---

**Supplement 1.**
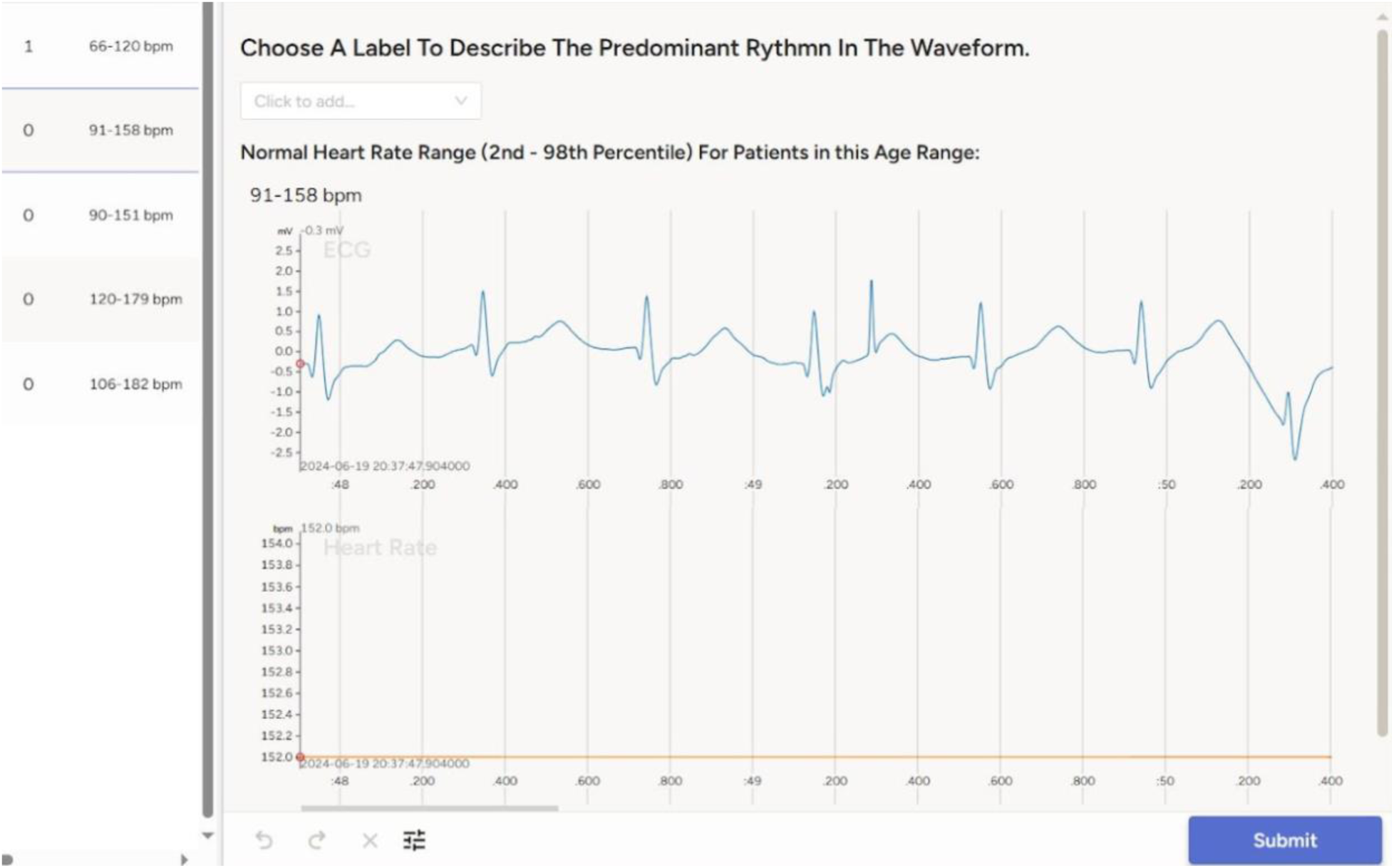
Example interface of Label Studio of discrete labeling tasks for users. **Column (left):** labeling tasks, ranges included are calculated normal ranges based on age **Row (top):** 10-second telemetry lead (II) task, oriented by time (x-axis) and voltage (y-axis) **Row (bottom):** heart rate variability across 10-second lead in the same labeling task

**Supplement 2**. PDF of Label Studio tutorial on navigating the interface

**Supplement 3.** PDF of REDcap survey for participants

